# Status of anemia and its associated factors in patients with Type 2 Diabetes Mellitus at a tertiary hospital in Nepal: An Observational Cross Sectional Study

**DOI:** 10.1101/2025.09.16.25335898

**Authors:** Suman Simkhada, Sarishma Ghimire, Mahesh Dhami, Pushpa Karki, Anuj Sharma Pandit

## Abstract

**Background:** Anemia is a common but under-recognized comorbidity among patients with type 2 diabetes mellitus (T2DM). Data on its prevalence and determinants in Nepal are limited. This study assessed the prevalence of anemia and its associated factors among T2DM patients attending a tertiary hospital.

**Methods:** A cross-sectional study was conducted in the Endocrinology Outpatient Department of KIST Medical College, Lalitpur, Nepal, from October 15, 2024, to January 15, 2025. The required sample size was 380, calculated using Cochran’s formula; 342 consecutive adults with T2DM on anti-diabetic medication were enrolled (90% of the planned 380, with negligible impact on precision). Data were obtained from structured interviews, medical records, and laboratory reports. Anemia was defined using WHO criteria. Bivariate associations were examined using the Chi-square test (crosstabulation). Multivariate binary logistic regression was used to identify independent factors and sensitivity analysis was conducted to confirm robustness of findings; model fit was assessed with Hosmer-Lemeshow test.

**Results:** Of 342 participants, 73 (21.3%) had anemia, with higher prevalence in females (22.9%) than males (19.6%). Most cases were mild (75.3%). Anemia prevalence increased with longer duration of diabetes and was more frequent in vegetarians, hypertensive patients, and those with thyroid disorders. In multivariate analysis, thyroid disorder (AOR=2.579, 95% CI: 1.307–5.086, p=0.006), longer diabetes duration (AOR=1.004 per month, 95% CI: 1.001–1.008, p=0.007), vegetarian diet (AOR=2.830, 95% CI: 1.286–6.228, p=0.01) and hypertension (p=0.008) were independently associated with anemia.

**Conclusion:** Anemia affected approximately one-fifth of T2DM patients, predominantly of mild severity. Thyroid disorders, longer disease duration, vegetarian diet, and hypertension were significant predictors. Routine screening and targeted management of anemia may improve outcomes in this population.

## INTRODUCTION

Diabetes Mellitus (DM) is a raised blood glucose either due to the body unable to produce enough amount or unable to use insulin effectively.[1] It can be classified as: type 1 DM, where destruction of pancreatic β-cells and absence of insulin, and type 2 DM, with insulin resistance.[2] Anemia is reduction of hemoglobin concentration in blood, reducing the oxygen-carrying capacity of RBC unable to meet the physiological requirements of body.[1] There is a significant association between hemoglobin and fasting blood glucose.

High incidence of anemia is likely to occur in patients with poorly controlled diabetes.[3] Anemia is unrecognized in 25% of diabetic patients.[1] Though high incidence, there is poor understanding of co-existence of anemia in diabetes especially in population of Southeast Asia.[4,5] Correction of the anemia among diabetics reduce complications and improves quality of life.[3]

Therefore, this study was designed to find out the Status of anemia and its associated factors in patients with type 2 Diabetes Mellitus at a tertiary hospital in Nepal.

## METHODS

### Study design and setting

This was a hospital-based observational, cross-sectional study was conducted at the endocrinology Out Patient Department (OPD) of KIST Medical College and Teaching Hospital, Lalitpur, Nepal, from 15th October 2024 to 15 January 2025. The study aimed to determine the prevalence of anemia and identify factors associated with it among adults with Type 2 DM.

### Participants

Adults aged >18 years with a confirmed diagnosis of Type 2DM and on anti-diabetic medication attending the OPD during the study period were eligible. Exclusion criteria included: Chronic Kidney Disease on hemodialysis, who had recent blood transfusion, pregnancy, severe critical illness, hematological malignancies or severe trauma. Participants were enrolled consecutively to reduce selection bias, as all eligible patients visiting the OPD during the study period were invited to participate.

### Sample size calculation and power

Sample size was calculated using Cochran’s Formula:

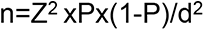

Where:

n = sample size
Z = Z-score = 1.96 (95% confidence level) P = Estimated proportion or prevalence
d = Desired precision= 0.05(5% margin of error)

Taking the pooled prevalence 45% from Meta-analysis by Mazumder et al in 2023 in South east Asia (P)= 0.45 [6] Sample size (n) = 1.96^2^ x 0.45 x (1-0.45)/0.05^2^

Calculated sample size(n) will be 380.32 ∼380. The calculated sample size was 380. Due to study period, 342 participants were enrolled representing 90% of the target.

### Standard guidelines for diagnosis

Anemia is when the diabetic patient’s Hemoglobin level was <13.0g/dL for a male or <12.0g/dL for a female. The severity was further classified as mild (11.0–12.9 for male, 11.0–11.9g/dl for female), moderate (8.0–10.9g/dl for both), and severe (<8.0g/dl for both) according to WHO.[1] Diabetes Mellitus Patient is when the patient was diagnosed as a Type 2 DM patient by a senior doctor and started anti-diabetic medication before data collection.[1] Glycemic control is labeled as good for the fasting blood sugar between 80 and 130 mg/dl (HbA1c<7%), and poor for <80 mg/dl or ≥130 mg/dl (HbA1c>7%).[7]

**Variable:** Variables used in this study were anemic status and sociodemographic variables, lifestyle factors, clinical and disease related factors.

*Dependent variable:* anemia status categorised as anemic and non anemic

#### Independent variables

Sociodemographic variable(age, gender, marital status, ethnicity, address, occupation, Education, religion)

Lifestyle factors (smoking status, alcohol intake, diet)

Clinical and disease related factors(Duration of Type 2 Diabetes Mellitus, Presence of Hypertension, Dyslipidemia, Thyroid disorder and complications, Glycemic control)

**Data sources/ measurement:** Data was collected by investigators on a daily basis using proforma, reviewing Lab reports & OPD cards. Laboratory personnel were blinded to study objectives.

**Bias:** Most participants were on Metformin, which can cause vitamin B 12 deficiency anemia. This was acknowledged as a potential confounder.

**Statistical Analysis:** Data were coded, entered and analysed using Statistical Package for Social Sciences (SPSS) software Version 21. Descriptive analysis included mean±SD for continuous variables and frequencies (%) for categorical variables. Bivariate associations were examined using the Chi-square test (crosstabulation). Multivariate binary logistic regression was used to identify independent factors and model fit was assessed with Hosmer-Lemeshow test. Statistical significance was set at p<0.05 with 95% Confidence Interval.

### Outcome Measures

Primary outcome measures: prevalence and severity of anemia Secondary outcome measures:

1. Sociodemographic characteristics (age, gender, marital status, ethnicity, address, occupation, Education, religion)
2. Lifestyle factors (smoking status, alcohol intake, diet)
3. Clinical and disease related factors (Duration of Type 2 Diabetes Mellitus, Presence of Hypertension, Dyslipidemia, Thyroid disorder and complications, Glycemic control)

## RESULTS

Of the 342 participants, 163 (47.7%) were male and 179 (52.3%) were female. Anemia was detected in 73 (21.3%) participants: 32 (19.6%) males and 41 (22.9%) females. The response rate was 100%. The socio-demographic and clinical characteristics are shown in *table 1 [Sociodemographic and clinical characteristics of Participants (n=342)]*.

**Table 1:** Socioden1ographic and clinical characteristics of Participants (n=342)

| Variable | Category | n(%) or Mean+-SD |
| --- | --- | --- |
| Age (years) | - | 55.6+-12.1 |
| Gender | Male | 163(47.7) |
|  | Female | 179(52.3) |
| Marital status | Married | 333(97.4) |
|  | unmarried | 9(2.6) |
| Address(Geographical) | Mountain | 18(5.3) |
|  | Hill | 264(77.2) |
|  | Terai | 60(17.5) |
| Religion | Hindu | 302(88.3) |
|  | Buddhist | 18(5.3) |
|  | Others | 22(6.4) |
| Ethnicity | Brahmin and Chhetri | 162(47.4%) |
|  | Janajati | 115(33.6) |
|  | Madhesi | 27(7.9) |
|  | Dalit | 26(7.6) |
|  | Others | 12(3.5) |
| Education level | None | 102(29.8) |
|  | Primary | 79(79) |
|  | Secondary | 107(31.3) |
|  | Tertiary | 54(15.8) |
| Occupation | Service | 91(26.6) |
|  | Business | 54(15.8) |
|  | Laborer and Farmer | 38(11.1) |
|  | Housewife | 127(37.1) |
|  | Others | 32(9.4) |
| Smoking history | Yes | 78(22.8) |
|  | No | 264(77.2) |
| Alcohol Consumption | Yes | 60(17.5) |
|  | No | 282(82.5) |
| Diet Type | Veg diet | 41(12.0) |
|  | Non-Veg diet | 301(88.0) |
| Duration of T2DM(months) | - | 103.8+-98.5 |
| HbA1c (%) | - | 8.1+-2.1 |
| Hb (gm/dL) | - | 13.8+-1.9 |
| Hypertension | Yes | 178(52) |
|  | No | 164(48) |
| Dyslipidemia | Yes | 264(77.2) |
|  | No | 78(22.8) |
| Thyroid Disorder | Yes | 75(21.9) |
|  | No | 267(78.1) |
| Complications of T2DM | Yes | 88(25.7) |
|  | No | 254(74.3) |

*Figure 1[Severity of anemia in patient with Type 2 DM]* shows the severity of anemia in patients with Type 2 DM: 55(75.3%) had mild anemia, 16(21.9%) had moderate anemia and 2(2.7%) had severe anemia. The comparative distribution of anemia severity by thyroid disorder, glycemic control, dietary habit and duration of Type 2 DM are illustrated in the figures 2-5 [Figure 2 *(Severity of anemia in patients with T2DM and thyroid disorder),* Figure 3 *(Glycemic control and severity of anemia),* Figure 4 *(Severity according to dietary habit),* Figure 5 *(Severity of anemia with duration of diagnosis of T2DM)*] respectively.

**Figure 1:**
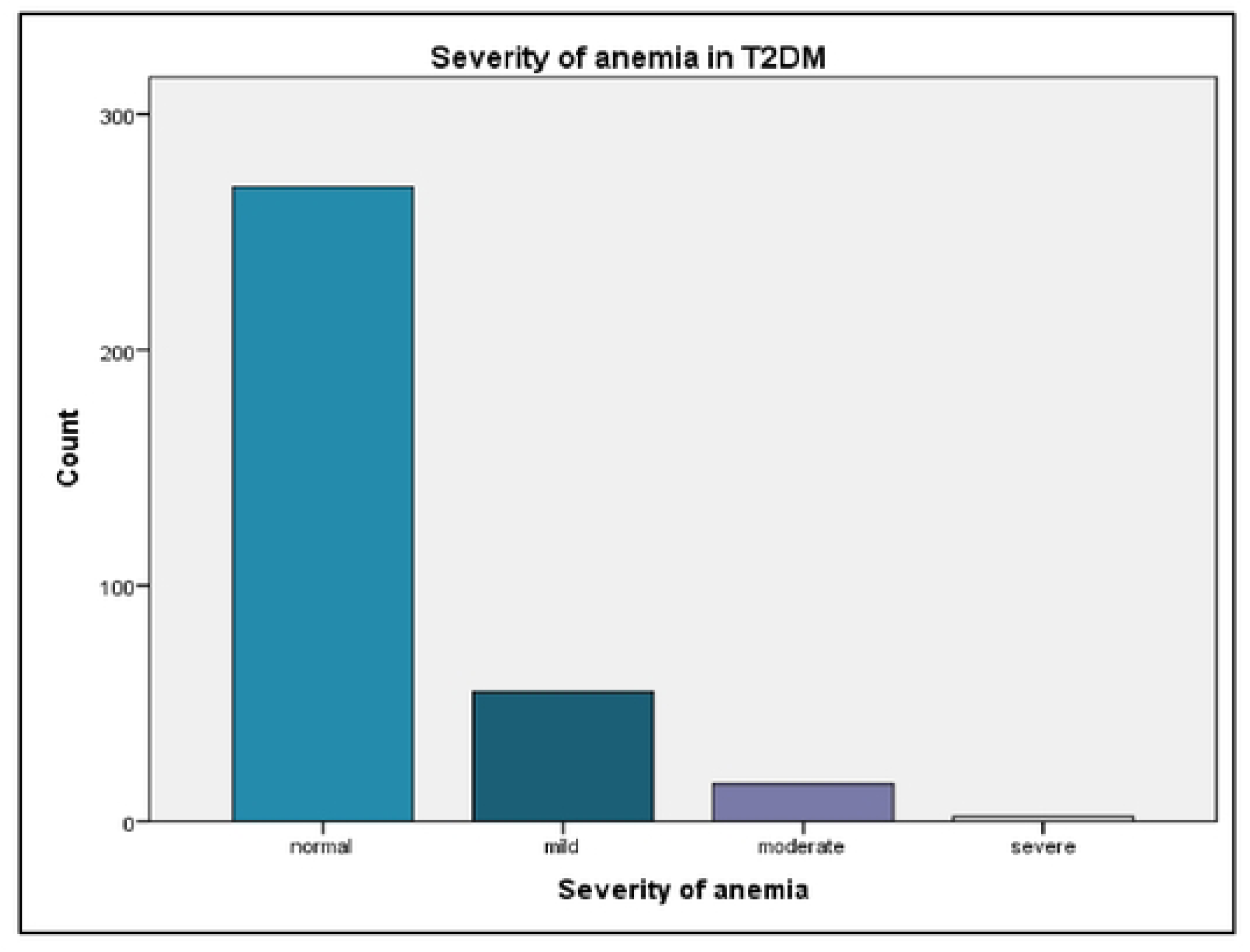
S**e**verity **of anemia in patient with Type 2 OM**

**Figure 2:**
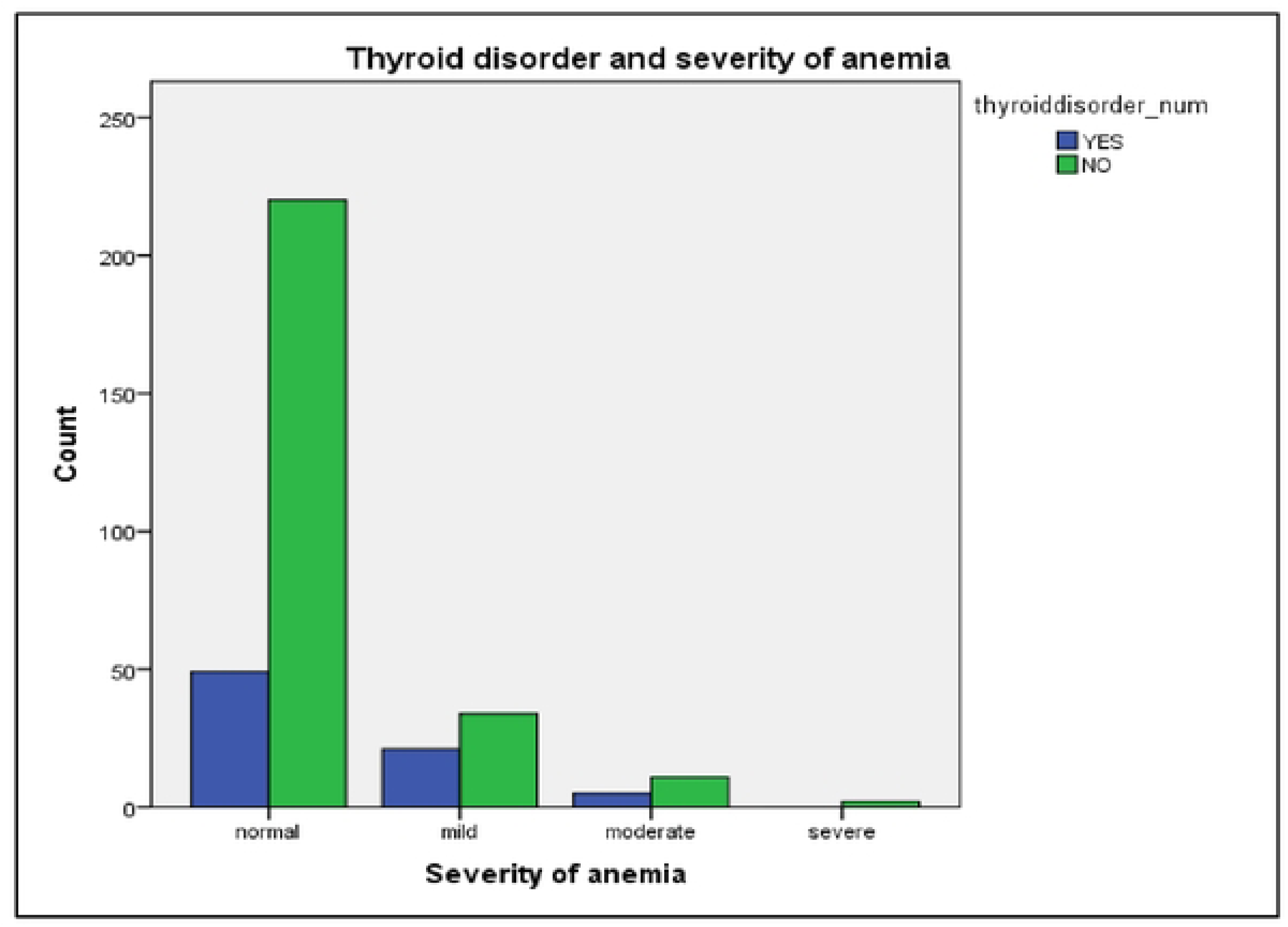
Severity of anemia in patients with T2DM and thyroid disorder

**Figure 3:**
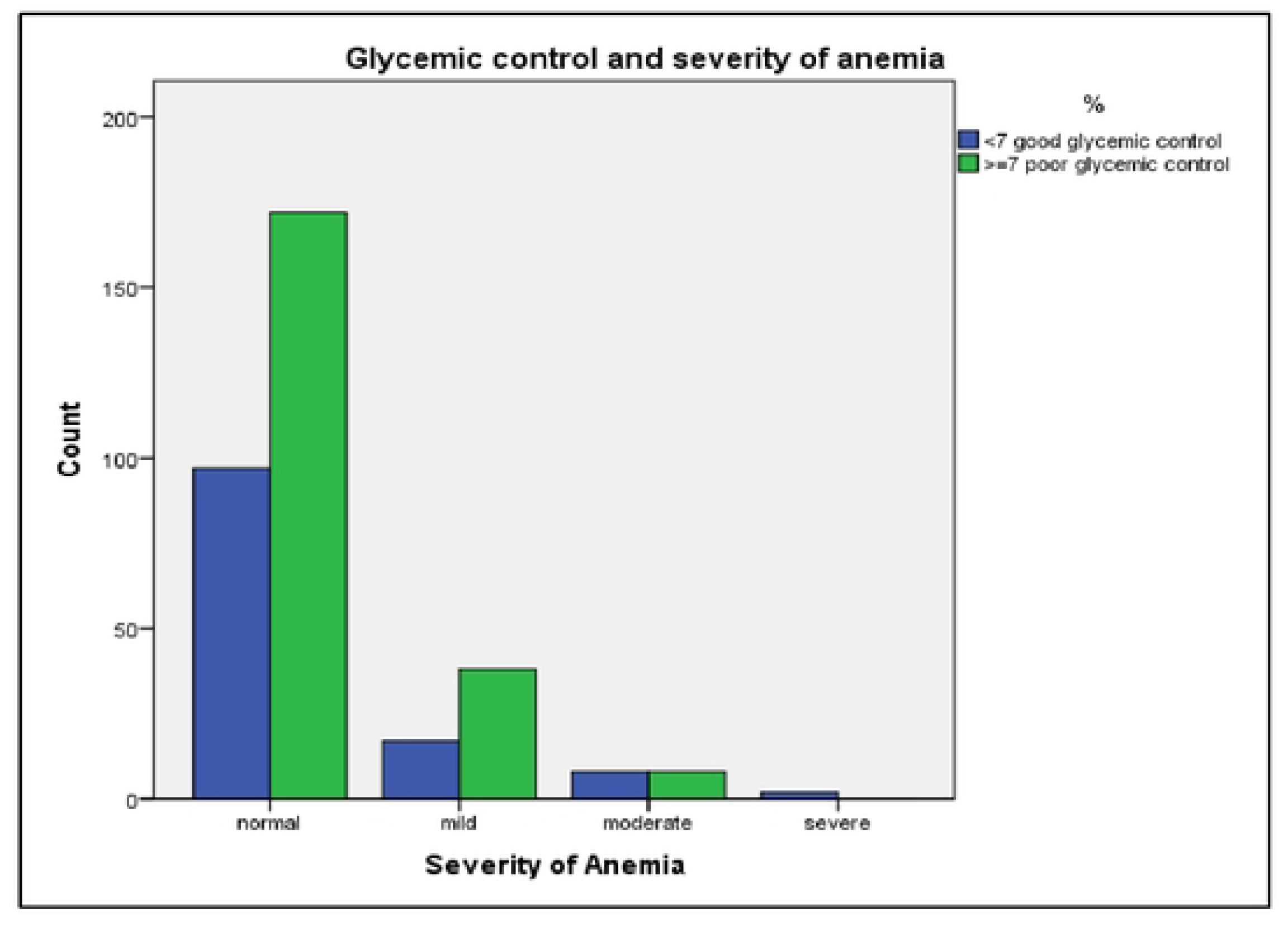
G**l**ycemic **control and severity of anemia**

**Figure 4:**
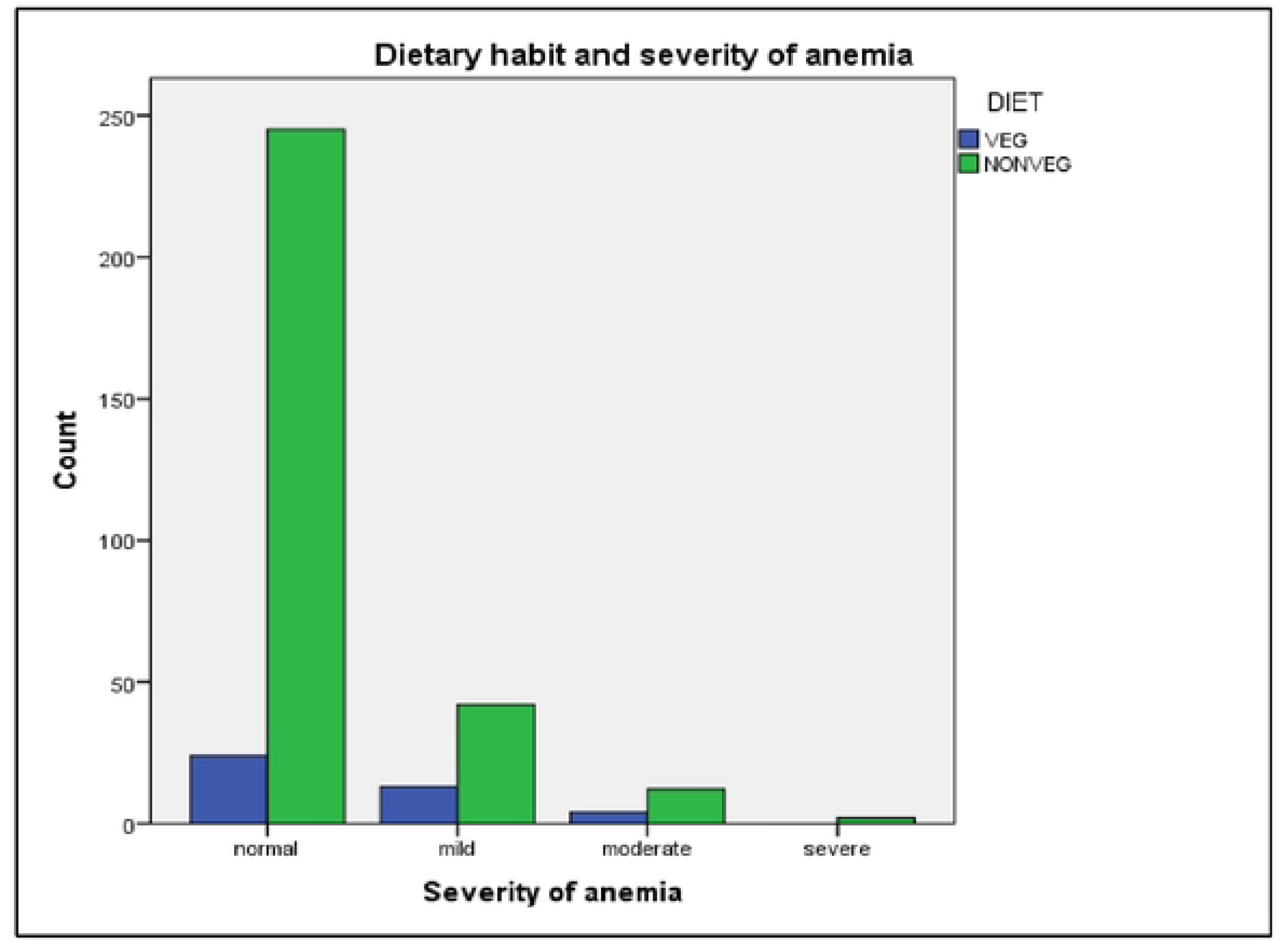
S**e**verity **according to dietary habit**

**Figure 5:**
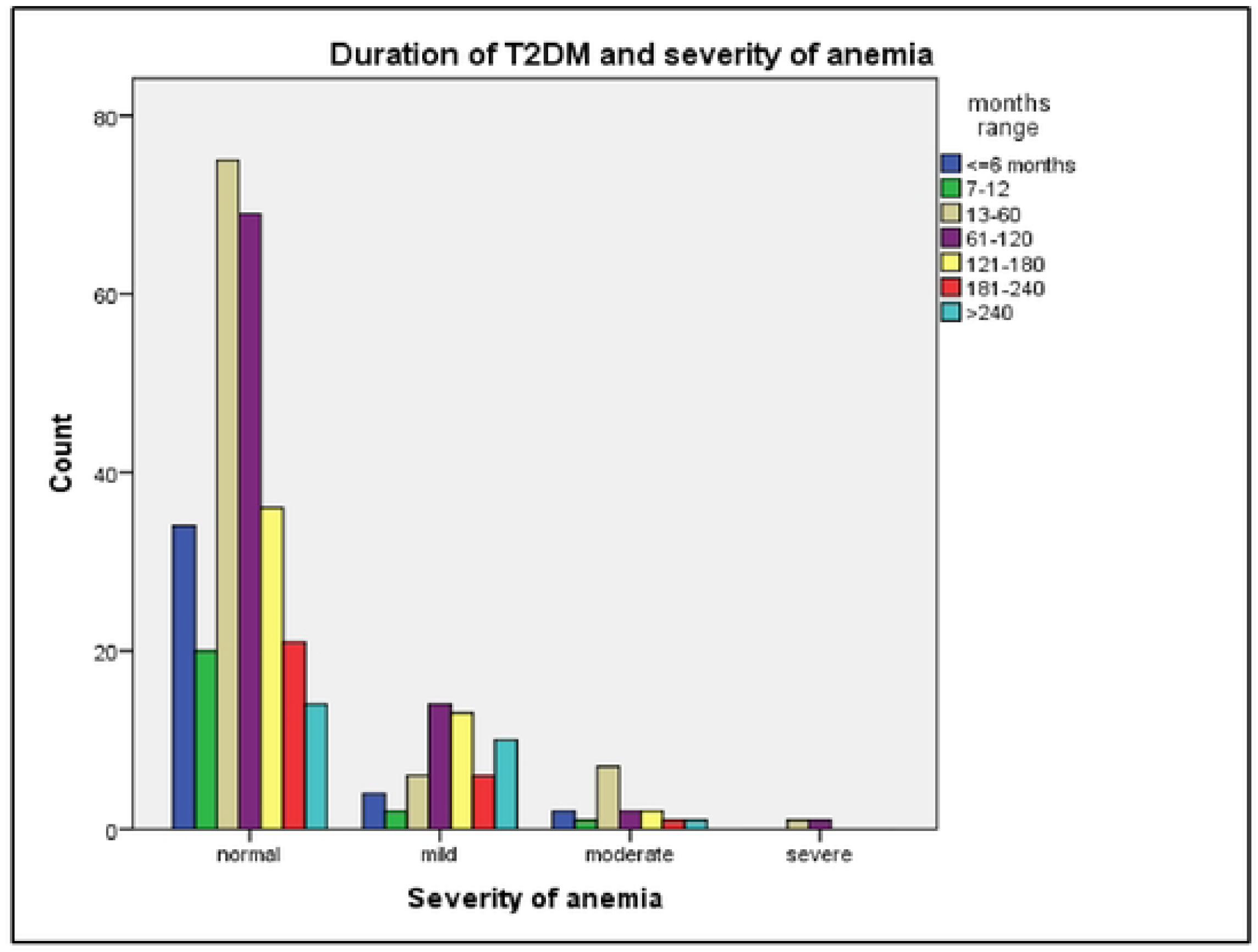
Severity of anemia with duration of diagnosis of T2DM

*Table 2* compares characteristics between anemic and non-anemic participants. The prevalence of anemia was highest among the participants >60 years of age. Anemia was observed in 46(21.1%) individuals with poor glycemic control and 27(21.7%) with good glycemic control. Prevalence increased with longer duration of Type 2DM, reaching 44% among those with disease duration >20 years, followed by 10-15 years and 15- 20 years.

**Table 2:** Comparisons of Characteristicsbetween anemic and Non-Anemic Participants (n=342)

| **Variables** | **Anemic n(%)** | **Non-Anemic n(%)** | **Chi-square value** | **df** | **p-value** |
| --- | --- | --- | --- | --- | --- |
| Age (range in years)<br>15-30<br>31-45<br>46-60<br>>60 | 1(12.5)<br>8(13.6)<br>33(20.5)<br>31(27.2) | 7(87.25)<br>51(86.4)<br>128(79.5)<br>83(72.8) | 4.894 | 3 | 0.180 |
| Gender<br>Male<br>female | 32(19.6)<br>41(22.9) | 131(80.4)<br>138(77.1) | 0.544 | 1 | 0.461 |
| Marital status<br>Married<br>unmarried | 70(21.0)<br>3(33.3) | 263(79.0)<br>6(66.7) | 0.791 | 1 | 0.374 |
| Address (Geographical)<br>Mountain<br>Hill<br>Teraí | 1(5.6)<br>57(21.6)<br>15(25.0) | 17(94.4)<br>207(78.4)<br>45(75.0) | 3.160 | 2 | 0.206 |
| Religion<br>Hindu<br>Buddhist<br>Others | 64(21.2)<br>5(27.8)<br>4(18.2) | 238(78.8)<br>13(72.2)<br>18(81.8) | 0.579 | 2 | 0.749 |
| Ethnicity<br>Brahmin and chhetri<br>Janajati<br>Madhesi<br>Dalit<br>Others | 32(19.8)<br>26(22.6)<br>8(29.6)<br>2(7.7)<br>5(41.7) | 130(80.2)<br>89(77.4)<br>19(70.4)<br>54(92.3)<br>7(58.3) | 7.296 | 4 | 0.121 |
| Education level<br>None | 26(25.5) | 76(74.5) | 2.478 | 3 | 0.480 |
| • Primary<br>• Secondary<br>• Tertiary | 18(22.8)<br>18(16.8)<br>11(20.4) | 61(77.2)<br>89(83.2)<br>43(79.6) |  |  |  |
| Occupation |  |  | 0.686 | 4 | 0.953 |
| • Service<br>• Business<br>• Laborer and farmer<br>• Housewife<br>• Others | 18(19.8)<br>11(20.4)<br>8(21.1)<br>30(23.6)<br>6(18.8) | 73(80.2)<br>43(79.6)<br>30(78.9)<br>97(76.4)<br>26(81.3) |  |  |  |
| Smoking history |  |  | 2.832 | 1 | 0.092 |
| • Yes<br>• No | 22(28.2)<br>51(19.3) | 56(71.8)<br>213(80.7) |  |  |  |
| Alcohol Consumption |  |  | 0.171 | 1 | 0.679 |
| • Yes<br>• No | 14(23.3)<br>59(20.9) | 46(76.7)<br>223(79.1) |  |  |  |
| Diet Type |  |  | 11.231 | 1 | 0.001 |
| • Veg<br>• Non-veg | 17(41.5)<br>56(18.6) | 24(58.5)<br>245(81.4) |  |  |  |
| Duration of T2DM(range in months) |  |  | 13.54 | 6 | 0.035 |
| • <=6 months<br>• 7-12<br>• 13-60<br>• 61-120<br>• 121-180<br>• 181-240<br>• >240 | 6(15.0)<br>3(13.0)<br>14(15.7)<br>17(19.8)<br>15(29.4)<br>7(25.0)<br>11(44.0) | 34(85.0)<br>20(87.0)<br>75(84.3)<br>69(80.2)<br>36(70.6)<br>21(75.0)<br>14(56.0) |  |  |  |
| HbA1c (%) glycemic control category |  |  | 0.021 | 1 | 0.884 |
| • <7(good)<br>• >=7(poor ) | 27(21.8)<br>46(21.1) | 97(78.2)<br>172(78.9) |  |  |  |
| Hypertension |  |  | **6.986** | **1** | **0.008** |
| • Yes | 48(27.0) | 130(73.0) |  |  |  |
| • No | 25(15.2) | 139(84.8) |  |  |  |
| Dyslipidemia |  |  | 0.547 | 1 | 0.460 |
| • Yes | 54(20.5) | 210(79.5) |  |  |  |
| • No | 19(24.4) | 59(75.6) |  |  |  |
| Thyroid Disorder |  |  | **10.155** | **1** | **0.001** |
| • Yes | 26(34.7) | 49(65.3) |  |  |  |
| • No | 47(17.6) | 220(82.4) |  |  |  |
| Complications of T2DM |  |  | 3.522 | 1 | 0.061 |
| • Yes | 25(28.4) | 63(71.6) |  |  |  |
| • No | 48(18.9) | 209(81.1) |  |  |  |

Values are presented as number(row% within respective group)

**Table 3.**
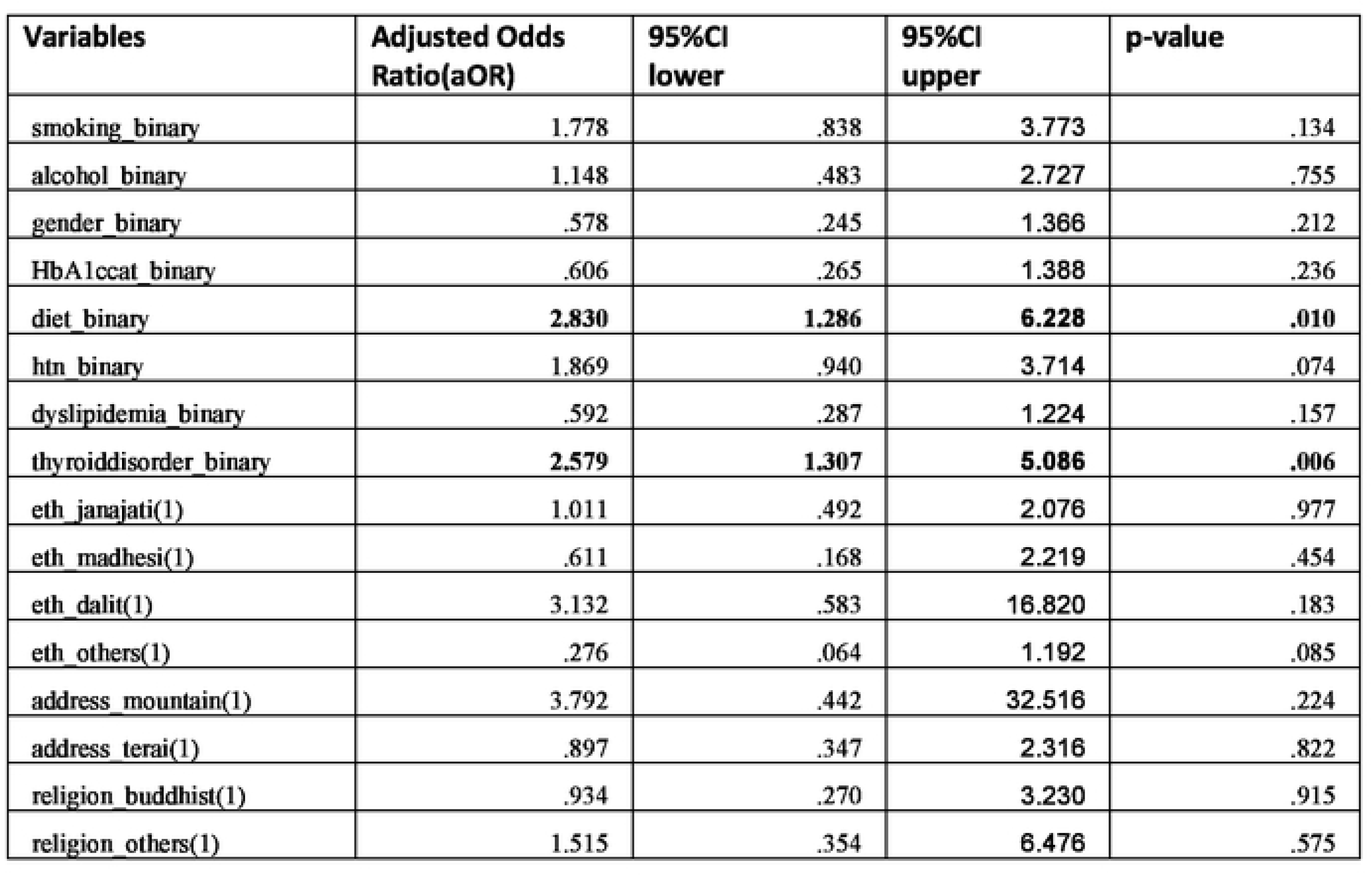
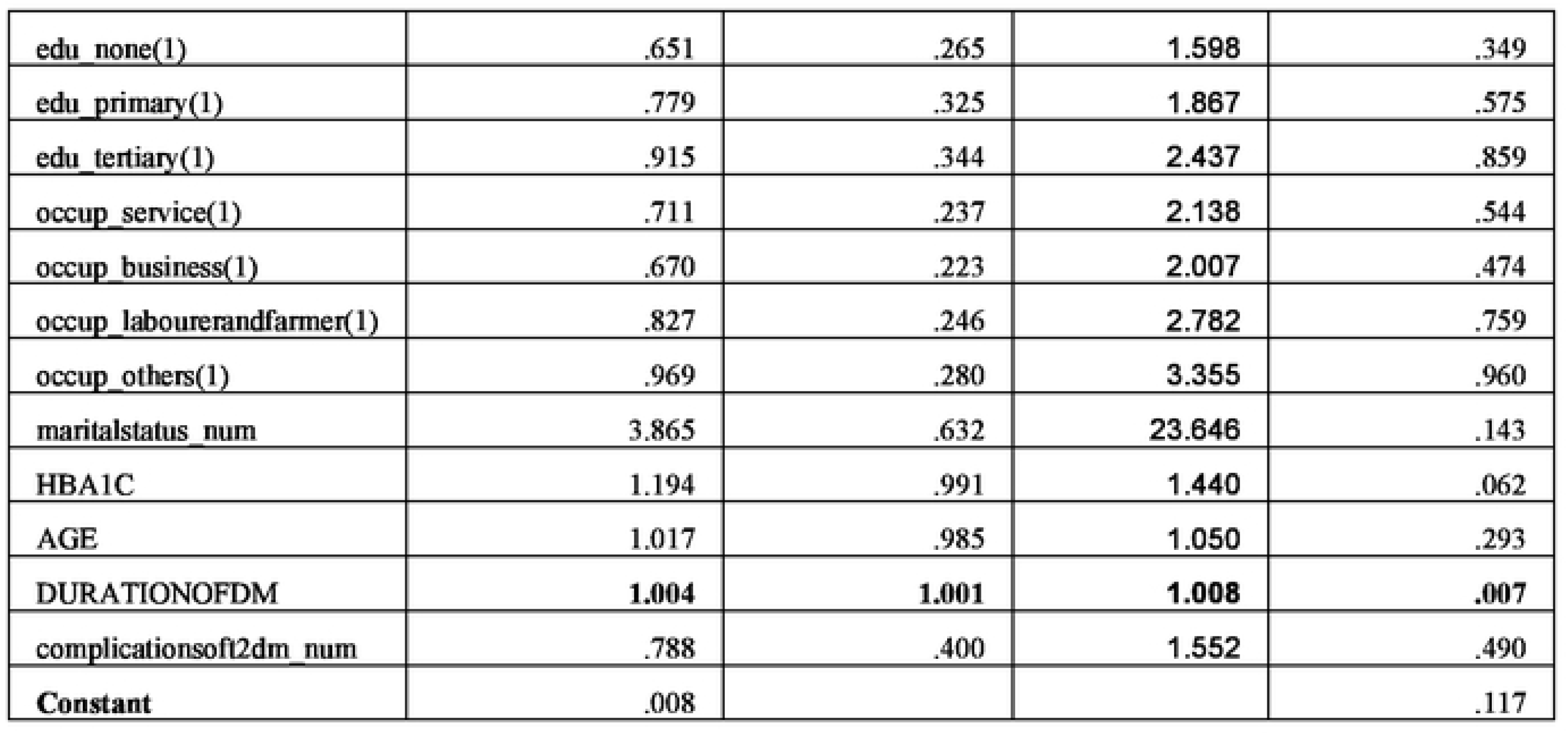
Multivariate Binary Logistic Regression Analysis of factors Associated with Anemia in Patients with T2DM (n=342)

| Variables | Adjusted Odds Ratio(aOR) | 95%CI lower | 95%CI upper | p-value |
| --- | --- | --- | --- | --- |
| smoking_binary | 1.778 | .838 | 3.773 | .134 |
| alcohol_binary | 1.148 | .483 | 2.727 | .755 |
| gender_binary | .578 | .245 | 1.366 | .212 |
| HbA1ccat_binary | .606 | .265 | 1.388 | .236 |
| diet_binary | **2.830** | **1.286** | **6.228** | **.010** |
| htn_binary | 1.869 | .940 | 3.714 | .074 |
| dyslipidemia_binary | .592 | .287 | 1.224 | .157 |
| thyroiddisorder_binary | **2.579** | **1.307** | **5.086** | **.006** |
| eth_janajati(1) | 1.011 | .492 | 2.076 | .977 |
| eth_madhesi(1) | .611 | .168 | 2.219 | .454 |
| eth_dalit(1) | 3.132 | .583 | 16.820 | .183 |
| eth_others(1) | .276 | .064 | 1.192 | .085 |
| address_mountain(1) | 3.792 | .442 | 32.516 | .224 |
| address_terai(1) | .897 | .347 | 2.316 | .822 |
| religion_buddhist(1) | .934 | .270 | 3.230 | .915 |
| religion_others(1) | 1.515 | .354 | 6.476 | .575 |
| edu_none(1) | .651 | .265 | 1.598 | .349 |
| edu_primary(1) | .779 | .325 | 1.867 | .575 |
| edu_tertiary(1) | .915 | .344 | 2.437 | .859 |
| occup_service(1) | .711 | .237 | 2.138 | .544 |
| occup_business(1) | .670 | .223 | 2.007 | .474 |
| occup_labourerandfarmer(1) | .827 | .246 | 2.782 | .759 |
| occup_others(1) | .969 | .280 | 3.355 | .960 |
| maritalstatus_num | 3.865 | .632 | 23.646 | .143 |
| HBA1C | 1.194 | .991 | 1.440 | .062 |
| AGE | 1.017 | .985 | 1.050 | .293 |
| DURATIONOFDM | **1.004** | **1.001** | **1.008** | **.007** |
| complicationsoft2dm_num | .788 | .400 | 1.552 | .490 |
| **Constant** | .008 |  |  | .117 |

Lifestyle factors also influenced prevalence: anemia was more common in participants consuming alcohol (14, 23.3%) and in smokers (22, 28.2%) compared to non-users. Vegetarian participants had a significantly higher prevalence of anemia (17, 41.5%) than non-vegetarian (56, 18.6%). Among 88(25.7%) participants with diabetes related complications, 25(28.4%) were anemic. Dyslipidemia 264(77.2%) was the most common comorbidity, followed by Hypertension 178(52%).

Bivariate analysis using the Chi-square test (crosstabulation) demonstrated significant associations between anemia and vegetarian diet (p=0.001), thyroid disorder (p=0.001), hypertension (p= 0.008) and increasing duration of Type 2 DM (p=0.035) as shown in the *Table 2 [Comparisons of Characteristics between anemic and Non-Anemic Participants (n=342)]*

Multivariate binary logistic regression analysis including all variables, with model fit assessed by the Hosmer-Lemeshow test as shown in *table no. 3 [Multivariate Binary Logistic Regression Analysis of factors Associated with Anemia in Patients with Type 2 DM (n=342)]* identified, thyroid disorder (AOR=2.579, 95%CI=1.307-5.086, p=0.006), longer duration of Type 2 DM (AOR=1.004, 95%CI=1.001-1.008, p =0.007) and vegetarian diet (AOR=2.830, 95%CI=1.286-6.228, p=0.010) as independent factors associated with anemia. Sensitivity analysis using a reduced model with clinically relevant predictors yielded consistent results, supporting the robustness of the findings. Overall, 342 participants were enrolled, representing 90% of the planned sample, with minimal impact on statistical precision.

## DISCUSSION

The prevalence of anemia among patient with Type 2DM in our study was 21.3% (73/342) with mild anemia being the predominant type (75.3%). Females were more frequently affected (22.9%) than males (19.6%) and anemia prevalence increased with age, particularly in participants over 60 years (27.2%). The median duration of Type 2 DM was 7 years, ranging from less than 6 months to more than 20 years, with longer duration associated with higher prevalence of anemia. Mean age of participants are 55.6 years (Standard Deviation = 12.1), which ranges between 20–95 years. Higher prevalence was seen with comorbid conditions like thyroid disorder, hypertension, dyslipidemia, vegetarian diet, with diabetes related complications; along with smoking and alcohol consuming habits. There mechanism of anemia in patients with type 2 DM remains incompletely understood. Studies suggest abnormal iron storage, albuminuria, hyperglycemia and neuropathy may increase the risk.[8]

### Prevalence of anemia and Comparison with other studies

The prevalence of anemia among patient with Type 2DM in our study was 21.3% (73/342) with mild anemia being the predominant type (75.3%). In one meta-analysis done by Mazumder et al among 14,194 participants with DM in south Asia, the pooled prevalence of anemia was 45% [6], higher than in our study. Another study by Ebben et al also showed significant occurrence of anemia in diabetic population.[9] Various studies revealed the development of anemia in T2DM patients is significantly associated with sex, age, marital status, educational status, BMI, hypertension, hematological diseases, glycemic control, gastrointestinal disorders, and chronic kidney diseases.[7]

### Sex and Age Differences in Anemia Prevalence

Anemia was more prevalent in females (22.9%) compared males (19.6%). This aligns with findings from Engidaw MT et al in 2020 and Mazumder et al where prevalence in females (48%) was more than males (39%); though contrasts with Kebede SA et al (10.56% males and 6.52% females) and Bekele A et al (males 41.5% and females 28.8%) reporting higher prevalence in males.[1,3, 6,7] The observed sex differences may be due to strict dietary habits of females to prevent weight gain, periodic menstrual bleeding, use of hormonal contraceptives etc.[10,11]

Age showed a clear association with anemia prevalence. Anemia was found to be more prevalent in the age group of more than 60 yrs 31(27.19%) and lowest in the age group 15 to 30 years 1(12.5%). This trend mirrors findings from Bekele A et al, Michalak et al and Mazumder et al.[4,6,7] It could be due to age-related decline in bone marrow function. It is documented that bone marrow cellularity decreases by 30% to 50% in individuals over 60 years of age, impairing hematopoiesis.[12]

### Glycemic Control and Diabetes Duration

For good glycemic control, fasting blood glucose measurement taken was <=130 mg/dL or (HbA1c<7%), and for poor glycemic control it was >130 mg/dl or (HbA1c>7%).[3,7] Of the total participants, 124(36.3%) had good glycemic control and 218(63.7%) had poor control. Anemia was observed in 46(21.1%) people having poor glycemic control and 27(21.7%) in people with good control, indicating no significant association between glycemic control and anemia (p= 0.884). This contrasts with the study by Bekele A et al which reported a 1.98-fold higher prevalence in poorly controlled diabetes .[7]

In contrast, duration of type 2DM showed a significant correlation with prevalence of anemia. In participants with a disease duration of ≤6 months, anemia prevalence was 15.0%, while it was 13.0% for 7–12 months and 15.7% for 13–60 months. This increased to 19.8% among those with 5–10 years (61–120 months) of diabetes. Notably, anemia prevalence rose to 29.4% in the 10–15 years group (121–180 months), 25% in the 15– 20 years group (181–240 months), and reached a peak of 44% in those with more than 20 years (>240 months) of diabetes. These results align with findings from studies in Northwest Ethiopia, Australia, India and Korea.[21,22,23,24] This could be due to the chronic effects of diabetes related hyperglycemia causing chronic hypoxia in the renal interstitium that causes change in vascular architecture, atypical cell growth, collagen proliferation and peritubular fibroblasts, which finally impaired erythropoietin production. Decreased erythropoietin causes reduction in RBC production.[25] In addition, A study by Redondo-Bermejo et al reported that red cell survival decreased by about 13% in hyperglycemic states.[26]

### Severity of anemia, Lifestyle and Dietary Factors

Considering the severity of anemia, mild anemia (75.3%) was the predominant type of anemia among patients with T2DM followed by moderate anemia 16(21.9%) in this study. This is in contrast with study done in Southern Ethiopia. This might be due to systemic inflammation, inhibition of erythropoietin release, rise in pro-inflammatory cytokines, and damage to the renal parenchyma.[13]

This study showed anemia was more prevalent in diabetic people consuming alcohol (23.3%). This was in line with the study conducted in southern Ethiopia, University of Washington Medical Center, Seattle, and New York.[13] It could be due to alcohol consumption that can cause micronutrient deficiency including iron and folate interfering with their absorption.[13,14] Furthermore it causes generalized suppression of blood cell production and the production of structurally abnormal blood cell precursors leading to premature destruction and anemia.[13]

This study showed significant association between dietary habit and anemia (AOR=2.830, 95%CI=1.286-6.228, p=0.010). Vegetarian diet consuming individuals (17,41.5%) were more affected, nearly 2-fold than non-vegetarians (56,18.6%). This finding was in line with study done in Malaysia, United States, and Saudi Arabia.[15,16,17] Exclusion of animal derived products from food can cause deficiency of micronutrients like Vitamin B12, iron, calcium, zinc, vitamin A etc.[18] A study showed Erythrocytes, hemoglobin, hematocrit, and serum ferritin were lower in vegetarians and vegans than omnivores,[19] As vitamin B12 is essential for the synthesis of nucleic acids, erythrocytes and iron for synthesis of hemoglobin their deficiency contributes significantly in development of anemia.[20]

### Diabetes-related Complications and Anemia

Of the total participants, 88(25.7%) people were living with complications of Type 2 DM, among which 25 (28.4%) people developed anemia. A study showed there was a statistically significant relation between diabetes related complications and the occurrence of anemia.[25] Also, anemia in patients with Type 2 Diabetes Mellitus (T2DM) is associated with a spectrum of complications that can significantly impact morbidity and mortality. The coexistence of anemia in T2DM may exacerbate diabetic complications and contribute to poorer clinical outcomes. One of the major complications linked to anemia in T2DM is diabetic retinopathy seen in 82(93.2%) of people living with complications of which 23(28.04%) developed anemia. Anemia has been implicated in the worsening of retinal hypoxia, potentially accelerating the progression of retinopathy in diabetic individuals.[27] Another critical concern is diabetic nephropathy. Anemia seen in 20(25.6%) of people having diabetic nephropathy. Anemia tends to appear early in diabetic kidney disease, often before significant reductions in glomerular filtration rate are evident.[28] Reduced erythropoietin production due to kidney damage in diabetes leads to normocytic normochromic anemia, which in turn worsens renal hypoxia and accelerates nephropathy progression.[29] Though our study doesn’t assess cardiovascular complications, several other studies show cardiovascular complications are notably more common in diabetic patients with anemia. The synergistic effect of diabetes-induced endothelial dysfunction and anemia-induced hypoxia can aggravate left ventricular hypertrophy, ischemic heart disease, and heart failure.[30] Studies have shown that anemic diabetic patients are at a higher risk of hospitalization for heart failure and have higher all-cause mortality.[31] These findings underscore the importance of routinely screening for and managing anemia in patients with Type 2 DM. Early detection and treatment may play a critical role in mitigating the risk of these complications and improving long-term outcomes.

### Comorbid Conditions Associated with Anemia

Dyslipidemia was the most common condition associated with Type 2 DM, seen in 264(77.2%) of the population. This is due to the fact that insulin resistance also affects lipid metabolism causing elevated Free Fatty Acid levels, chylomicrons and lipoproteins in the circulation.[32] This finding is lower than the study by Junaid et al (97.9%) in elderly population and Ogbera et al (89.1%) in age groups similar to our study. But this is higher than the study that reported 70.5% in Eritrea among an elderly population.[33,34,35]

Hypertension was the next commonly associated condition and one of the strongest risk factors for development of anemia.[3] Of total, 178(52%) were hypertensive and 48 (65.8%) of them developed anemia. It could be due to Hypertension along with hyperglycemia together accelerates renal impairment thus increasing the risk. [3]

Thyroid disorder was observed in 75(21.9%) with significant association with anemia (AOR=2.579, 95%CI=1.307-5.086, p=0.006)which is more than the study done in Civil Hospital Karachi, Pakistan by Syeda Iffat Bukhari et al.[36] Among them, anemia was seen in 26(34.7%) which is similar to the study done by Saif Aboud M Alqahtani in Saudi Arabia.[37] Anemia in hypothyroidism might result from bone marrow depression, decreased erythropoietin production, comorbid diseases, or concomitant iron, vitamin B12, or folate deficiency.[38] Thus, coexistence of hypothyroidism and diabetes increases the risk of developing anemia.

### Strengths and Limitations

A strength of this study lies in its comprehensive inclusion of clinical, demographic, and behavioral factors, supported by robust sample size (n = 342). Although enrollment reached 90% of the planned sample size, the impact on statistical precision was minimal, as reflected by narrow confidence intervals for prevalence estimates and consistent results in sensitivity analysis. Glycemic control was assessed using HbA1c which is a more reliable and standardized indicator of long-term glucose control compared to random or fasting blood sugar levels.

As a cross-sectional study, causal relationship between anemia and associated factors could not be established. Our study was conducted in one center that may limit its generalizability. This did not include clinical assessment of diseases like chronic kidney diseases, clinical evaluation of anemia. Besides this, Smoking and alcohol data were collected in binary format (Yes/No) which limited assessment of quantity, frequency and duration of exposure. Diabetes duration was grouped into clinically meaningful intervals based on disease course and population distribution; while the intervals are unequal, they reflect relevant clinical stages and ensure adequate group sizes for analysis

## CONCLUSION

Anemia affects approximately one-fifth of patients with type 2 DM attending this tertiary hospital in Nepal, with the majority of cases being mild. Independent predictors of anemia include thyroid disorders, longer duration of diabetes, vegetarian diet and hypertension. These findings highlight the importance of routine anemia screening and targeted management of type 2 DM patients to prevent potential complications and improve long-term clinical outcomes.

## Data Availability

All the data files are available from the zenodo database (accession number 10.5281/zenodo.17011162 .

## Data availability Statement

The data used to support the findings of this study are included within this article. *All the data files are available from the zenodo database (accession number* **10.5281/zenodo.17011162 .**

## Ethical Approval

This study was approved by the Institutional Review Committee, KIST Medical College and Teaching Hospital (Reference number: 2080/81/98). Then, permission letters from officials of KIST Medical College and Teaching Hospital, Department of Endocrinology, were processed before data collection. Written Informed Consent was obtained from all the participants. To ensure confidentiality, patient names were not included; instead, code numbers were assigned to depict the results.

## Authors Contribution

Conceptualization: SG, SS Methodology: SS, MD Software: ASP, MD, SG Formal Analysis: MD, SG

Investigation: SS, SG, MD, PK, ASP Writing-Original Draft: SG, MD, PK, ASP

Writing-Review and Editing: SG, MD, SS, PK Supervision: SS

SS: Suman Simkhada SG: Sarishma Ghimire MD: Mahesh Dhami PK: Pushpa Karki

ASP: Anuj Sharma Pandit

All authors read and approved the final manuscript.

## ACKNOWLEDGMENTS

The authors would like to express our sincere gratitude to study participants for their willingness to participate in this study. The authors also express their appreciation to KIST Medical college and teaching hospital and staff of DM clinic for support in data collection throughout this study. Also, we’d like to thank Institutional Review Committee (IRC), KIST Medical College and Teaching Hospital for their invaluable support and granting permission to carry out this research and Biostatistician, Associate Professor Prem Prasad Panta for his guidance during data analysis.

## ACRONYMS/ ABBREVATIONS

AOR: Adjusted Odds Ratio ß -cells: Beta cells
CI: Confidence Interval
CKD: Chronic Kidney Disease df: Degree of freedom
DM: Diabetes Mellitus Gm/dl: Gram / deciliter Hb: Hemoglobin
HbA1C: Glycated Hemoglobin C
OPD: Out Patient Department p: Probability Value
RBC: Red Blood Cells
SD:: Standard Deviation
WHO: World Health Organization

